# Prevalence and Determinant of Adverse Drug Reactions Among MDR-TB Patients Attending St. Peter’s TB Specialized Hospital, Addis Ababa, Ethiopia

**DOI:** 10.1101/2022.08.03.22278306

**Authors:** Fikru Letose Shararo, Degemu Sahlu Asebe, Hawine Teshome Teshome, Sahle Asfaw Jabo

**Affiliations:** Gambella Peoples’ Regional National State Health Bureau, Gambella, Ethiopia; College of health Science, department of public health, Selale University, Fiche, Ethiopia; College of health science, Addis Ababa University, Addis Ababa, Ethiopia; College of health Science, Kotbe metropolitan University, Addis Ababa, Ethiopia

**Keywords:** Adverse drug reaction, MDR-TB, Institution based cross sectional study

## Abstract

**Background:** Tuberculosis is one of a major public health problem throughout the world. About 9.6 million people were estimated to have Tuberculosis in 2014. Out of this, 480 000 cases were multidrug-resistant Tuberculosis. Thus, it has to be identified, focused and prioritized for subsequent and targeted interventions. Therefore, the aim this study was to assess the prevalence and factors affecting adverse drug reaction among multidrug-resistant Tuberculosis patients.

**Method:** Institution based retrospective cross sectional study design was included all patients with Multi Drug Resistant who were registered and treated from 9 January 2020 to 30 March 2020. Data abstraction form was used to obtain patient information from their cards. Data were collected by trained nurses who have been working at multidrug-resistant Tuberculosis treatment service. Data were entered, cleaned and checked using Epi Data version 4.6 statistical software and analyzed by SPSS version 23. Bivariate and multivariate logistic regression analyses were used to examine the association between independent and dependent variables. The results were presented as Odds Ratio with 95% CI.

**Result:** A total of 286 patients were included in the study. The prevalence of adverse drug reaction in this study were 169 (59.1%). Factors such as Treatment outcome and comorbidity were significantly associated with adverse drug reaction among multidrug-resistant Tuberculosis patients.

**Conclusion and Recommendation:** There was high prevalence of adverse drug reaction among multidrug-resistant Tuberculosis patients. Comorbidity and treatment outcome were the independent determinants. Patients who are identified with adverse drug events need special attention enhanced clinical management.

## Introduction

Tuberculosis (TB) is an infectious disease caused by Mycobacterium tuberculosis; a rod-shaped bacillus called “acid-fast” due to its staining characteristics in laboratory. Occasionally the disease can also be caused by Mycobacterium Ovis and Mycobacterium africanum(1).

TB is considered drug-resistant when the TB causative agent is not killed by one or more of the available anti-TB drugs. Drug-resistant TB can be primary or secondary (acquired). Primary resistance is drug resistance among new cases, while secondary resistance is drug resistance among previously treated cases(1, 2).

Treatment for multidrug-resistant Tuberculosis (MDR-TB) is longer, and requires more expensive and more toxic drugs. For most patients with MDR-TB, the current regimens recommended by WHO lasts 20 months and treatment success rates are much lower(3). In Ethiopia, MDR-TB patients are treated with standardized second line treatment regimen for at least 18-24 months. Drug Resistance Survey (DRS) data from representative patient populations are used to design a standardized treatment regimen(2).

Globally, an estimated 3.3% of new TB cases and 20% of previously treated cases have MDR-TB in 2014. From these MDR-TB cases, an estimated 190,000 people died. Despite the number of TB deaths is unacceptably high, with a timely diagnosis and correct treatment almost all people with TB can be cured(3).

The prevalence and mortality of Tuberculosis of all forms is estimated to be 546 and 73 per 100,000 populations respectively. In the year 2006/7 Ethiopia registered 129,743 cases of TB. According to latest estimates, Ethiopia stands 7^th^ in the list of High Burden Countries for TB.

According to the MOH hospital statistics data, tuberculosis is the leading cause of morbidity, the third cause of hospital admission (after deliveries and malaria), and the second cause of death in Ethiopia, after malaria(1).

Patients with MDR-TB take more tablets and receive more injections for a longer period of time, may experience more adverse effects and require increased support to continue treatment and/or to monitor adverse effects. Detecting and controlling adverse effects in a timely manner promotes adherence and prevents default to treatment(1, 4).

Unmanaged adverse reactions greatly affect the patients’ adherence to treatment regimen, which in turn leads to the development of drug resistance(4). This study is intended to determine the current management practice of adverse drug reactions among MDR-TB patients on second-line anti-tuberculosis drug regimen. In addition, health care practitioners’ knowledge and patient’s understanding of their treatments will be assessed with this study.

## Material and Methods

### Study design, setting, and period

Institution based retrospective cross sectional study was conducted at St. Peter’s TB Specialized Hospital from 9 January 2020 to 30 March 2020.

It was inaugurated on February 04, 1963 as “St. Peter’s TB Clinic” by Emperor Haile Selassie I. St. Peter’s has served as the only specialised TB referral hospital in Ethiopia for four decades. Afterwards, by the government’s decision TB treatment centres were opened in every region of the country and that has alleviated the burden which had been laid on St. Peter’s. St. Peter’s is the first hospital to offer medical services to patients with drug resistant TB. The hospital has a total of 400 staffs during the study period and 286 patients with MDR-TB who were registered and treated during 9 January 2020 to 30 March 2020.

### Study participants and sampling procedures

All MDR-TB patients who were treated with anti-TB drugs at St. Peter’s TB Specialized Hospital were the source population. Whereas, all MDR-TB patients attended St. Peter’s TB Specialized Hospital and treated by second-line anti-TB drugs and aged 18 and above were study population. All records of patients who have been on treatment from 9 January 2020 to 30 March 2020 at St. Peter’s TB Specialized Hospital were reviewed (**Fig 1**) and 286 patients profile cards that fulfill the inclusion criteria were incorporated in the study.

**Figure 1.**
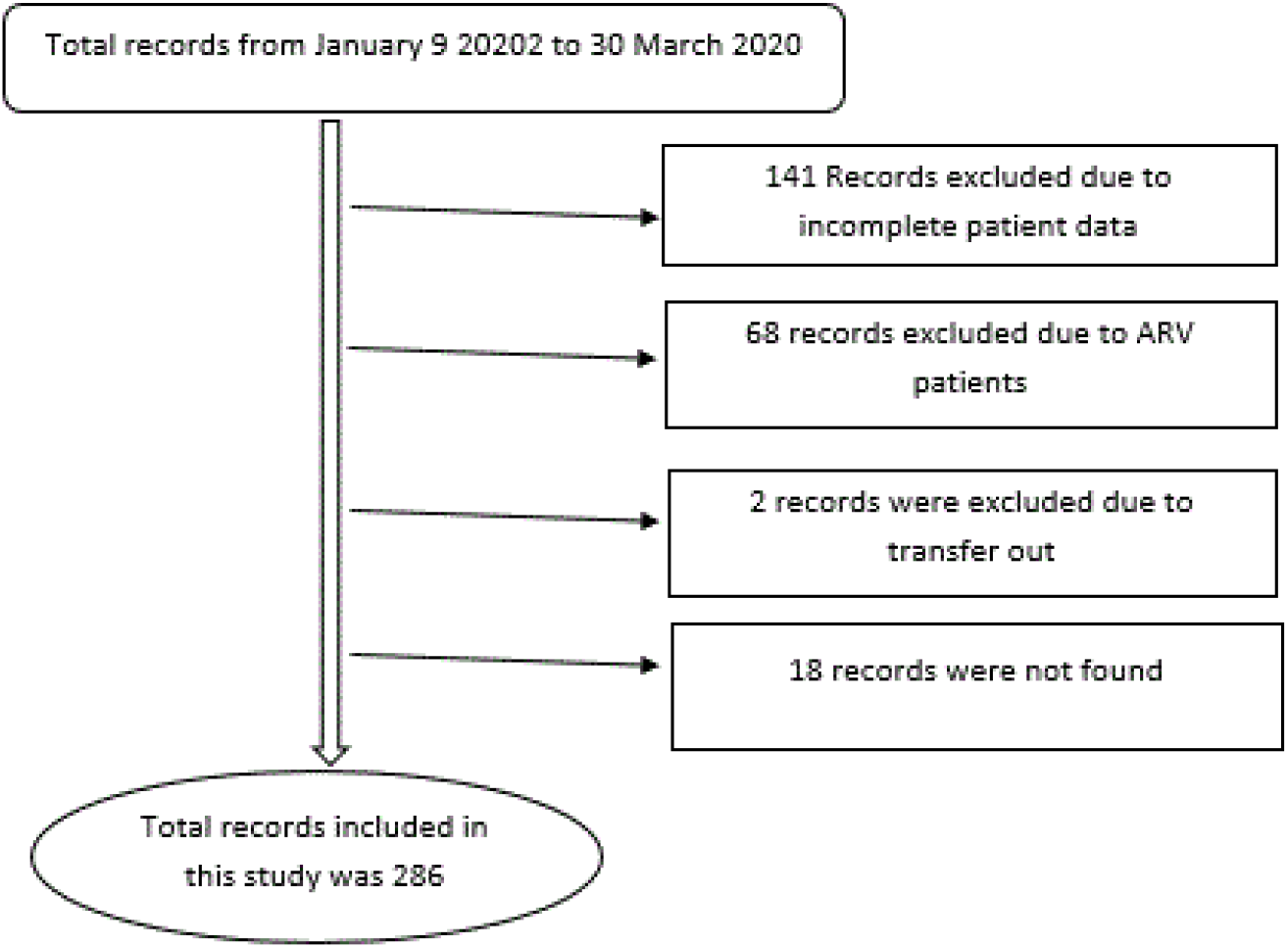
Schematic representation of selection of patient profile cards among the patients attending St. Peter hospital, Addis Ababa, 2009 – 2016.

### Data collection instruments and methods

A data abstraction form was developed so as to meet the objectives of this study. It was used to obtain the required information from the patient’s medical chart. The form consisted of questions regarding socio-demographic characteristics, clinical characteristics, treatment regimens, adverse drug reactions and ADR managements. The list of patients needed for the study was obtained from the hospital’s database. After getting the list of the patients, their charts were retrieved from the record and documentation office. A data abstraction format was used to record the necessary information from patients’ chart. The data were collected by trained nurses who are staff of MDR-TB ward.

### Variables of the Study

The dependent variable was adverse drug reactions. Whereas, the independent variables include Age, Sex, Marital status, educational status, Occupational status, Co-morbidity, MI, Site of TB, Anti-TB drugs, Treatment outcome, History of smoking, History of alcohol use and Drug abuse history.

### Measurements

#### New

A patient who has received no or less than one month of anti-tuberculosis treatment.

#### Relapse

A patient who was previously treated for TB and whose most recent treatment outcome was “cured” or “treatment completed”, and who is subsequently diagnosed with recurrent episode of TB.

#### Treatment after being lost to follow-up

A patient after taking treatment for more than one month who returns to treatment following interruption of treatment for two or more consecutive months.

#### Treatment after failure of New TB regimen

A patient who has received new regimen for TB and in whom treatment has failed. Failure is defined as sputum smear/culture positive at five months or later during treatment.

#### Treatment after failure of Retreatment regimen

A patient who has received retreatment regimen for TB and in whom treatment has failed. Failure is defined as sputum smear/culture positive at five months or later during treatment.

#### Transfer in

A patient who has been transferred from another TIC to continue MDR-TB treatment.

#### Other

Refers to any DR-TB patient who does not fit into any of the above categories.

#### Cured

Treatment completed according to national recommendation without evidence of failure and three or more consecutive cultures taken at least 30 days apart are negative after the intensive phase.

#### Treatment completed

Treatment completed according to national recommendation without evidence of failure but no record that three or more consecutive cultures taken at least 30 days apart are negative after the intensive phase.

#### Treatment failed

Treatment terminated or need for permanent regimen change of at least two anti-TB drugs because of; lack of conversion by the end of the intensive phase, or bacteriological reversion in the continuation phase after conversion to negative after intensive phase, or evidence of additional acquired resistance to fluoroquinolones or second line injectable drugs, or adverse drug reactions.

#### Lost To Follow Up (LFTU)

A patient whose treatment was interrupted for two consecutive months or more

#### Died

A patient who dies for any reason during the course of treatment

#### ADR

A response to a drug which is noxious and unintended, and which occurs at doses normally used in human for the prophylaxis, diagnosis, or therapy of disease, or for the modification of physiological function.

#### Side Effects

An expected, well-known reaction resulting in little or no change in patient management.

#### Data quality control

Four data collectors (Diploma nurse) were trained on the content of the questionnaire and the objective of the study for 03 days. A pre-test was done at 5% of the sample size, at another district before the actual data collection. A structured interviewer administered questionnaire was used for the data collection. The supervisor and the investigator were checked the data for completeness, missing, and unwanted filling daily. Epi data manager version 4.6.0.4 was used for data entry. SPSS version 23 was used for the analysis.

#### Ethical Consideration

Ethical clearance was obtained from the ethics Institutional Review Board (IRB) of Addis Ababa University. A formal letter from Addis Ababa University was submitted to St. Peter’s TB Specialized Hospital and permission was obtained. Written informed consent for participation was not required for this study. To ensure confidentiality, name and other identifiers of the patients were not recorded on the data abstraction formats.

#### Data processing and Analysis

Data was entered, cleaned and checked for the completeness using Epi data version 4.6 statistical software. Errors related to inconsistency were verified using cross data exploration techniques. Then managed data was exported to Statistical Package for Social Sciences (SPSS) software version 23. Descriptive analysis was done using frequency and percentages for the variables. Logistic regression was used to analyze the associations between different variables. Factors that were associated with ADR at 95% significance level in the bivariate analysis were included in the final multivariate analysis. Statistical significance was declared if the p-value is less than 0.05.

## Results

### Socio-Demographic Characteristics

The mean age (±SD) of the patients who have participated in this survey was 29.58 years (±11.4) and their age ranged from 18 to 75 years. Regarding the sex of the participants 159 (55.6%) were males. More than half, 181 (63.3%) of the patients were from Addis Ababa, followed by the Oromia region 45 (15.7%). Almost equal numbers of the patients are single 136 (47.6%) and married 138 (48.3%). Most of the patients have attended education within the grade levels 1 up to 8, 93 (32.5%) and 9 up to 12, 86 (30.1%) (**Table 1**).

**Table 1:** Socio-demographic characteristics of MDR-TB patients attending St. Peter hospital, Addis Ababa, 2020.

| Variable | Frequency | Percent |
| --- | --- | --- |
| Sex |  |  |
| Female | 127 | 44.4 |
| Male | 159 | 55.6 |
| Age group (years) |  |  |
| 18-27 | 166 | 58.3 |
| 28-37 | 68 | 23.9 |
| 38-47 | 26 | 8.8 |
| 48-57 | 11 | 3.9 |
| >58 | 15 | 5.3 |
| Marital status |  |  |
| Single | 136 | 47.6 |
| Married | 138 | 48.3 |
| Separated | 4 | 1.4 |
| Divorced | 8 | 2.8 |
| Region |  |  |
| Addis Ababa | 181 | 63.3 |
| Oromia | 45 | 15.7 |
| SNNPR | 24 | 8.4 |
| Others** | 36 | 12.4 |
| Educational status |  |  |
| Unable to read and write | 27 | 9.4 |
| Read and write | 30 | 10.5 |
| Grade 1-8 | 99 | 34.6 |
| Grade 9-12 | 86 | 30.1 |
| Certificate/ Diploma | 18 | 6.3 |
| Degree and above | 26 | 9.1 |
| Occupational status |  |  |
| Governmental employee | 30 | 10.5 |
| Non-governmental employee | 19 | 6.6 |
| Daily laborer | 17 | 5.9 |
| Self-employed | 75 | 26.2 |
| Unemployed | 18 | 6.3 |
| Student | 75 | 26.2 |
| Others*** | 52 | 18.2 |

### Clinical Characteristics

Most of the patients had pulmonary TB, 270 (94.4%). Additionally, majority of the patients; 244 (85.3%) had a prior history of TB treatment (failure, default and relapse cases). The majority of the patients 270 (94.4%) had no co-morbidities while the rest had co-morbidities such as diabetes and cardiomyopathy. The most frequently used 251 (87.8%) treatment regimen was capreomycin (Cm), levofloxacin (Lfx), pyrazinamide (Z), ethionamide (Eto)/Prothionamide (Pto) and cycloserin (Cs). (**Table2**)

**Table 2:** Clinical characteristics of MDR-TB patients attending St. Peter hospital, Addis Ababa, 2020.

| Characteristics | Frequency | Percent |
| --- | --- | --- |
| **Site TB** |  |  |
| PTB | 270 | 94.4 |
| EPTB | 16 | 5.6 |
| **Registration group** |  |  |
| New | 42 | 14.7 |
| Relapse | 43 | 15.0 |
| After lost to follow up | 6 | 2.1 |
| After failure of first treatment | 121 | 42.3 |
| After failure of re-treatment | 74 | 25.9 |
| **Previous second line drug use** |  |  |
| Yes | 11 | 3.85 |
| No | 275 | 96.15 |
| **Co-morbidities** |  |  |
| Yes | 17 | 5.90 |
| No | 269 | 94.10 |

| **Base-line BMI category** |  |  |
| --- | --- | --- |
| <18.5Kg/m2 | 186 | 65.03 |
| 18.5-24.9Kg/m2 | 97 | 33.92 |
| >=24.9 Kg/m2 | 3 | 1.05 |

### Frequency, management and Treatment outcomes of ADR

Out of 286 patients, 169 (59.1%) were found to be experiencing symptoms associated with ADRs. Most of the patients experienced combination of ADRs. The most common ADR was nausea and vomiting (57.4%) followed by dyspepsia and abdominal pain (44%) and arthralgia (43.5%). Other ADRs also included headache (22.5%), insomnia (21.3%), anorexia (18.9%), and others (**Table3)**. From 169 patients, proper management of ADR was given for 119(70.4%) patients **(Fig2**). One hundred fifty-three (53.5%) patients completed their treatments. While 78 (27.3%) of the patients were cured (**Table 3)**.

**Table 3:** Adverse drug reactions which occurred among the patients attending St. Peter hospital, Addis Ababa, 2020.

| **ADR** | **Frequency** | **Percent** |
| --- | --- | --- |
| Rash | 12 | 7.1 |
| Nausea and vomiting | 97 | 57.4 |
| Dyspepsia and abdominal pain | 74 | 44 |
| Diarrhea | 8 | 4.7 |
| Hepatitis | 3 | 1.8 |
| Arthralgia | 73 | 43.5 |
| Electrolyte abnormalities | 5 | 3 |
| Nephrotoxicity | 9 | 5.4 |
| Ototoxicity | 8 | 4.8 |
| Peripheral neuropathy | 13 | 7.7 |
| Depression | 12 | 7.1 |
| Headache | 38 | 22.5 |
| Psychosis | 12 | 7.1 |
| Seizures | 2 | 1.2 |
| Hypothyroidism | 2 | 1.2 |
| Anxiety | 11 | 6.5 |
| Insomnia | 36 | 21.3 |
| Visual disturbances | 14 | 8.3 |
| Musculoskeletal pain | 10 | 6 |
| Vestibular toxicity | 10 | 6 |
| Anorexia | 32 | 18.9 |
| Pain at the injection site | 5 | 3 |
| Others*** | 27 | 16 |

\*\*\*Fever, sweating, body weakness, anemia, weight loss, palpitation, fungal infection, vaginal discharge.

**Fig 2:**
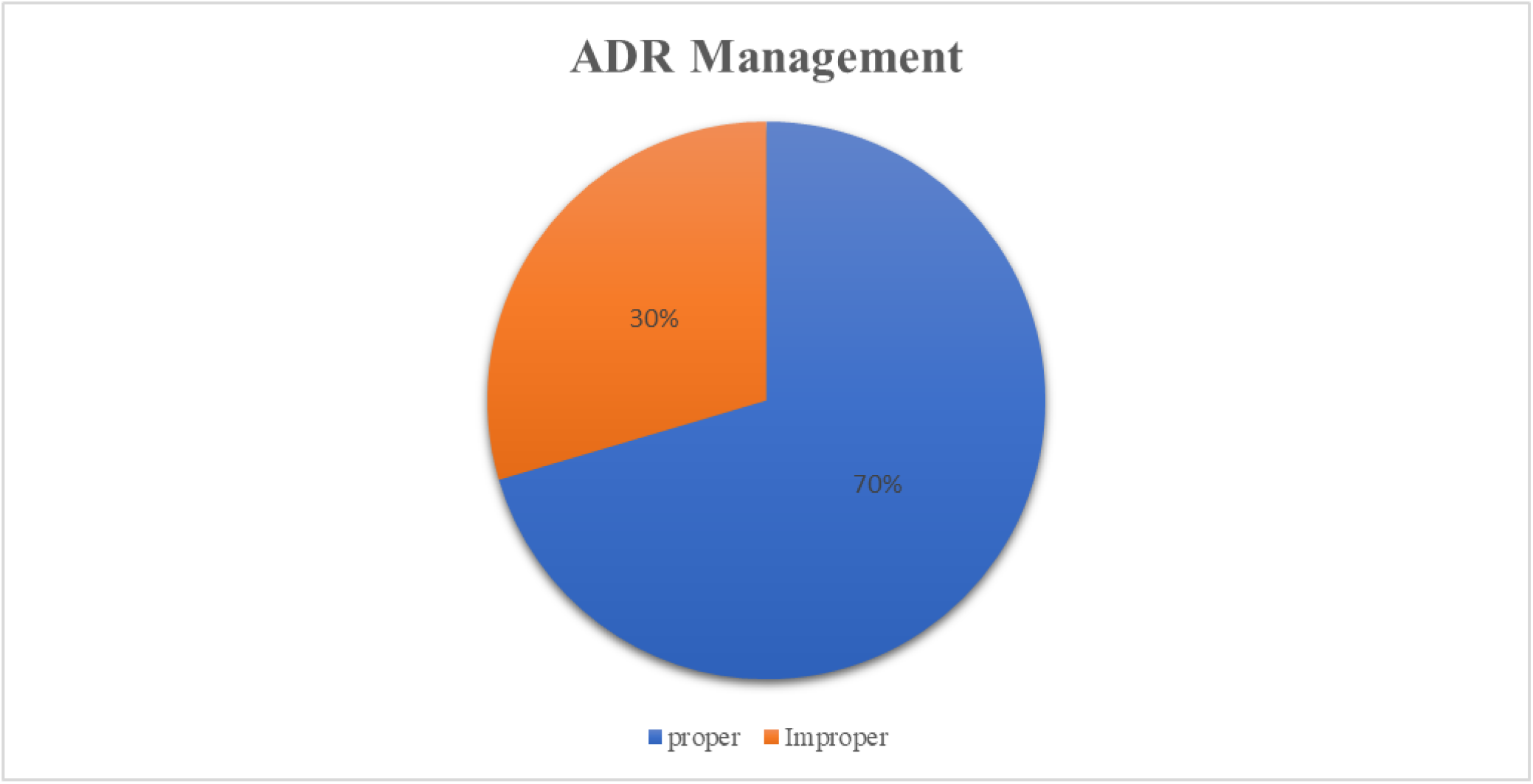
Management of ADR among the patients attending St. Peter hospital, Addis Ababa, 2009 – 2016.

### Treatment regimens used

The highest number 251 (87.8%) of the patients took the standard treatment regimen which is the combination of Cm, Lfx, Pto/ Eto, Cs, Z during the intensive phase and Lfx, Pto/ Eto, Cs, Z during the continuation phase (**Table 4**).

**Table 4:** Treatment outcomes and the most common anti-TB regimens used among patients in St. Peter hospital, Addis Ababa, 2020.

| **Treatment outcomes** | **Frequency** | **Percent** |
| --- | --- | --- |
| **Cured** | 78 | 27.3 |
| **Completed** | 153 | 53.5 |
| **Lost to follow up** | 23 | 8.0 |
| **Died** | 23 | 8.0 |
| **Failure** | 9 | 2.1 |
| **Regimen** |  |  |
| Cm, Lfx, Pto/ Eto, Cs, Z | 251 | 87.8 |
| Cm, Lfx, Pto/ Eto, Cs, PAS, Z | 12 | 4.2 |
| Cm, Lfx, E, Pto/ Eto, Cs, Z | 6 | 2.1 |

### Multivariate analysis of factors and adverse drug reaction

Variables with P <0.02 during the bivariate analysis were included in the multivariate logistic regression analysis to see the effect of confounding variables. The multivariate analysis indicated that comorbidity and treatment outcomes showed statistically significant association with adverse drug reaction. But age and smoking did not show significant association with adverse drug reaction. The odds of adverse drug reaction in patients who had comorbid diseases were about 6 times higher than those who had no the diseases (AOR = 5.809, 95%CI (1.106 - 30.504)). Treatment outcome was significantly associated with adverse drug reaction. Patients who had treatment outcome of cure were more likely to develop adverse drug reaction (AOR=10.012, 95% CI, (2.131 - 47.036)) than those who had treatment outcome of failure (**Table 5**)

**Table 5:** Multivariate analysis of factors for Adverse Drug Reaction among the patients attending St. Peter hospital, Addis Ababa, 2020.

| Variables | COR (95%CI) | P value | AOR (95%CI) | P value |
| --- | --- | --- | --- | --- |
| **Age group (years)** |  |  |  |  |
| 18-27 | 0.486(0.165 - 1.429) | 0.190 | 1.098(0.306 - 3.938) | 0.886 |
| 28-37 | 0.496(0.159 - 1.549) | 0.227 | 0.960(0.251 - 3.671) | 0.952 |
| 38-47 | 0.200(0.050 - 0.794) | 0.022 | 3.632(0.778 - 16.949) | 0.101 |
| 48-57 | 0.250(0.047 - 1.344) | 0.106 | 3.311(0.480 - 22.849) | 0.224 |
| >58 | 1 |  | 1 |  |
| **History of Cigarette Smoking** |  |  |  |  |
| Yes | 4.792(1.061 - 21.649) | 0.042 | 2.889(0.510 - 16.367) | .231 |
| No | 1 |  | 1 |  |
| **Co-morbidities** |  |  |  |  |
| Yes | 5.601(1.256 - 24.976) | 0.024 | 5.809(1.106 - 30.504) | **0.038** |
| No | 1 |  | 1 |  |
| **Treatment outcomes** |  |  |  |  |
| Cured | 0.104(0.024 - 0.462) | 0.003 | 10.012(2.131 - 47.036) | **0.004** |
| Complete | 0.730(0.189 - 2.823) | 0.648 | 1.367(0.334 - 5.601) | 0.664 |
| LTFU | 1.829(0.374 - 8.937) | 0.783 | 0.657(0.126 - 3.426) | 0.618 |
| Died | 1.829(0.374 - 8.937) | 0.456 | 0.366(0.064 - 2.083) | 0.257 |
| Failure | 1 |  | 1 |  |

## Discussion

In this study, 59.1 % of the participants developed symptoms associated with ADRs these is higher compared with Previous studies have documented a prevalence rate of 44% in Nigeria(5),50% in India(6),50.5% in Vietnam (7). Another study in India has also found a prevalence of 33.96%(8). This finding is considerably higher than those given in previous reports and the observed variation could be a result of disparity in the definitions of adverse drug reactions used by the researchers, the study design used, the geographic variation, the specific anti-TB drugs used and the time at which the study was conducted.

The most common ADR experienced by participants were nausea and vomiting (57.4%) followed by dyspepsia and abdominal pain (44%) and arthralgia (43.5%). Other reported ADRs include headache (22.5%), insomnia (21.3%) and anorexia (18.9%). The result is inconsistent with those reported by (4-8). This difference may be attributed to the difference in the pharmacokinetic parameter of the patients, use of different anti –TB regimens and their doses among countries.

The drugs most likely associated with nausea and/or vomiting were capreomycin, levofloxacin, ethionamide/protionamide and pyrazinamide. Most of the aforementioned drugs also cause dyspepsia. This finding is also comparable the other findings reported by Haregewoin and Petros (4, 9, 10).Majority of the patients experienced gastrointestinal symptoms during early weeks of treatment. These symptoms may prevent the delivery of adequate therapy and lead patients for further problem. Gastrointestinal side effects which were commonest can be largely prevented by proper timing and spacing of drugs with food and if necessary, giving antiemetic, antacids and proton pump inhibitors (PPIs) or H2 receptor blockers.

Arthralgia was found to be one of the common ADR encountered by patients in this study. It was likely caused by pyrazinamide and levofloxacin(9).The present finding is inconsistent with study findings from India(8).The discrepancy may be due to variation in inclusion of the causative drugs in the national MDR-TB treatment protocol between the countries.

This study identified determinants of adverse drug reaction among the patients attending St. Peter hospital in Addis Ababa Ethiopia. Comorbidity and treatment outcome were the independent determinants of adverse drug reaction of the patients. But it found that any of the socio demographic characteristics were not statistically significant with the development of adverse drug reaction.

Treatment outcome was significantly associated with adverse drug reaction. The odds of adverse drug reaction in patients whose treatment outcome of cure were about 10 times higher than (AOR=10.012, 95% CI, (2.131 - 47.036)) those whose treatment outcome of failure. The study finding was also in line with studies conducted in India and Russia (6, 10-12). The possible reason could be those patients who were cure might be closely followed by tuberculosis treatment care providers and as a result they got better reporting of ADRs and favorable treatment outcome.

The study revealed that MDR-TB patients who had comorbid diseases were about 6 times more likely (AOR = 5.809, 95%CI (1.106 - 30.504)) to develop adverse drug reaction than those who had not the disease. This finding is consistent with previous reports(13-15). In the presence of comorbidities, the patients take several drugs, whether prescription or over the-counter, contributes to the risk of having an ADR. The number and severity of ADRs increases disproportionately as the number of drugs taken increases. It may also be related to medications that are useful in the treatment of one disease may precipitate or worsen another condition.

This study showed that history of smoking, alcohol use, drug abuse and socio demographic characteristics such as sex, age, living condition, marital status, educational status and occupation of MDR-TB patients were not significantly associated with the development of adverse drug reactions. However, previous study has reported that age, sex, smoking and alcohol use had significant association with adverse drug reactions (4, 16-20). The possible explanation for this disparity might be the existence of variation with regard to genetic factors, culture, belief and living conditions. It might also be related to difference in the number of study participants used among the studies.

### Strengths and Limitations of the study

Clinical record reviewed using pre-tested structured data collection format which minimize information bias. Data quality was assured by trained nurse data collectors and close supervision.

The use of complete data from registers and patient cards, and for the reporting of adverse drug reactions and associated factors is also one the strength of this study. Whereas the Reporting bias might exist, as data were obtained retrospectively through medical card review, especially for adverse events not defined by laboratory tests, such as gastrointestinal disorders, which might overestimate or underestimate the magnitude of adverse events. Some important confounding factors might not have been collected in this study. Electronic medical records are designed for daily use in hospital clinical diagnosis and treatment, not for analysis of adverse reactions/events of drugs used, so the records might lack some information needed to detect adverse events, and these were likely to result in bias of the study.

### Conclusion and recommendation

During the study period, the prevalence of adverse drug reaction among MDR-TB patients was found to be higher. Majority of the study participants experienced more than one adverse drug reactions. Nausea, vomiting followed by dyspepsia and abdominal pain and arthralgia were the commonly reported ADRs.

The study showed that comorbidity and treatment outcomes were the independent determinants adverse drug reaction among MDR-TB patients in the hospital. However, socio-demographic variables and behavioral factors were not significantly associated with the development of ADRs

Based on the finding of this study, the following important recommendations are forwarded management strategies should be strengthened to minimize ADR occurrence and for better treatment outcome. MDR-TB Patients having comorbid diseases should be closely monitored and gave special counseling to minimize the development of ADRs. Patients should be educated on the possible ADRs of MDR-TB drugs. Further investigation should be performed emphasizing the occurrence of adverse events in different phases of MDR-TB treatment, and corresponding relationships with comorbidities, sociodemographic and behavioral variables.

### Disclosure

The authors reports no both financial and non -financial competing interest in these works

## Data Availability

All data produced in the present study are available upon reasonable request to the authors.

